# Emotional responses toward COVID-19: A longitudinal assessment of age differences

**DOI:** 10.1101/2021.03.21.21254050

**Authors:** Marta Malesza

## Abstract

The current study investigates the relation between age and emotional responses and coping strategies at two moments during the spread of COVID-19 in Poland, namely the first peak (March-May 2020) and the second pick (October-December 2020). A sample of 414 individuals between the ages of 18 and 81 were asked to rate the intensity of the ‘shock’, ‘sadness’, ‘anger’, and ‘fear’ they experienced due to COVID-19 and respond to items from the Brief Cope questionnaire. The present findings demonstrate that anger was consistently less intense among older adults than younger ones. Emotion-focused coping strategies were more commonly used by younger adults than middle-aged or older ones at the first peak of the outbreak; however, this trend had reversed during the second peak of the pandemic, as the older age groups demonstrated a far greater increase in the use of this form of coping. Results indicate a greater ability to use emotional regulation among older adults than younger ones, as the former are less likely to react to a crisis through anger and more able to adapt coping mechanisms to a dynamic environment.

## Introduction

The first case of the disease later named COVID-19 was seen in the Chinese city of Wuhan in December 2019, after which it spread rapidly beyond national borders. On March 11, 2020, the World Health Organization (WHO) declared a pandemic (Nishiura et al., 2020). By February 19, 2021, 1,623,218 Poles had been infected and there had been 41,823 fatalities (World Health Organization, 2021). Faced with this situation, intense feelings of anxiety, fear, and worry were frequently expressed, with many individuals demonstrating coping strategies including distancing and enhanced personal hygiene practices (Barber & Kim, 2021; Restubog, Ocampo, & Wang, 2020; Shanahan et al., 2020; Trnka & Lorencova, 2020).

### Age and emotional reactions

The emotional response of older adults to problems is different to that demonstrated by younger ones (Charles, Carstensen, & McFall, 2001; Labouvie-Vief & Medler, 2002). One study showed respondents pictures of a woman struggling in a nursing home and asked them to describe the feelings it triggered in them. Older adults were more likely to feel sadness while younger people tended towards anger (Charles et al., 2001). A second study similarly found less anger among older adults than younger ones when they were asked to react to negative situations (Weiner & Graham, 1989). Moreover, the results of laboratory studies such as the two described above have been reproduced by studies investigating real-life situations. For example, older adults are less likely than younger ones to display anger or hostility when faced with chronic illness. The same holds true of response to natural disaster: a study examining responses to the 1994 Northridge Earthquake found lower levels of distress among older people as well as a lower tendency to dwell on the risk of further earthquakes thereafter (Knight, Gatz, Heller, & Bengtson, 2000).

### Age and coping strategies

Different age groups do not only display different emotional reactions to problematic situations, but also tend to practice different coping strategies. ‘Coping’ can be understood as how individuals think and behave when managing stress, and two principal types of coping strategies have been determined (Folkman & Lazarus, 1980; Folkman, Lazarus, Pimley, & Novacek, 1987). The aim of *problem-focused coping* is to solve a problem or change some aspect of the problematic situation, while that of *emotion-focused coping* is to mitigate the emotional distress caused (Folkman & Lazarus, 1980). Prior research has shed light on the importance of domain specificity and cognitive appraisal as moderators in the relationship between coping strategies and age (Blanchard-Fields, Chen, & Norris, 1997; Charles et al., 2001).

Most studies of the relationship between age and choice of coping strategy have found that older people are more likely to use emotion-focused strategies and younger ones are more likely to use problem-focused ones. When asked to imagine experiencing difficulties in a nursing home, older adults tended to turn to non-action-oriented strategies, while younger ones were more likely to turn to action-oriented ones (Charles et al., 2001; Folkman et al., 1987; Watson & Blanchard-Fields, 1998). Similarly, when asked to consider how they would feel if the Three Mile Island nuclear generating facility were to be restarted, older adults living nearby were more likely to leverage emotion-focused coping strategies than their younger neighbors (Prince-Embury & Rooney, 1990).

### The relationship between emotions and coping

Prior research has amply demonstrated the association between age group and emotional response, as well as the likelihood that younger and older adults will adopt different types of coping strategy (Barber & Kim, 2021; Charles et al., 2001; Shanahan et al., 2020). However, to date little research has investigated the relationship between the two. There is no consensus in the literature about whether specific coping strategies derive from emotions or, on the contrary, that the emotions derive from the strategies (Fredrickson, 2001). It is thought that motivational elements within emotions can trigger certain behaviors or actions: experiencing fear, for example, will cause individuals to seek a means of escape and protect themselves, while experiencing anger can give rise to the wish to strike out, disgust engenders the wish to expel, and joy causes a desire to be playful (Charles et al., 2001). Furthermore, associations have been identified between emotions and specific coping strategies (Folkman et al., 1987). Adults who experience greater levels of anger and fear are more likely to employ active-oriented coping strategies, including talking about whatever event is triggering the emotion. Those who experience sadness, on the other hand, tend to employ non-active coping strategies, for example deciding to accept or avoid the difficult situation. Coping strategies, if properly deployed, should offer means for individuals to mitigate the stress caused by problematic situations and events and experience fewer negative emotions (Folkman & Lazarus, 1980). However, the trajectory of the association between emotions and coping strategies, like that of the association between emotions, coping strategies, and age groups, can only be identified by longitudinal studies, none of which have, to our knowledge, been carried out during the current COVID-19 pandemic (Barber & Kim, 2021; Restubog et al., 2020; Shanahan et al., 2020; Trnka & Lorencova, 2020). The present study attempts to bridge this gap by longitudinally investigating the association between emotions felt and coping strategies adopted in the face of COVID-19, as well as whether the association varies according to age group.

### Purposes of the present study

The current study aims to investigate how three adult-aged cohorts – younger, middle-aged, and older – in Poland experienced emotions and deployed coping strategies in the face of the COVID-19 outbreak. In so doing, the study contributes to research on the association between age difference and emotions, and age difference and coping strategies, as well as the relationship between emotions and coping strategies. The following two hypotheses were formulated on the basis of the literature review presented above: firstly, that older adults demonstrate less anger and fear (active emotions) in response to a crisis but more sadness (nonactive emotion) than younger ones; and secondly, that older adults deploy more emotion-focused coping strategies and fewer problem-focused ones to deal with the COVID-19 pandemic. Specific hypotheses in regard to the relationship between coping and emotions were not formulated. If emotions determine coping strategies, it was anticipated that differences in coping strategies toward COVID-19 can be accounted for by the differences in emotional response to the outbreak over time. If, on the other hand, coping determines subsequent emotions, the reverse proposition would hold true.

Furthermore, the present research tried to identify any differences in these relationships across age groups. Given that the pandemic is naturally occurring and has a universal impact, and noting the consequent limits on controllability, investigating the association between age and the emotional response and coping strategies deployed enabled us to ensure that predictability and controllability remained constant for all cohorts. Moreover, as the study was longitudinal, participants created their own controls; hence, controls for dispositional factors which might impact emotional response and/or coping behaviors were controlled for.

## Method

### Participants

Data were gathered in two phases. The first spanned the period 27^th^ March–14^th^ May, 2020, when the first pandemic peaked in Poland. A total of 697 respondents were recruited in total: 278 in the 18–35-year age group, 224 in the 36–59-year age group, and 195 in the ≥ 60 age group. Hence, respondents were grouped into three groups: younger (39.9%), middle-aged (32.1%), and older (28.0%). As regards gender, 68.3 percent were female (n = 476) while in terms of educational attainment, 8.3 percent had completed primary school (n = 58), 40.2 percent had completed secondary school (n = 280), and the remaining 51.5 percent had undertaken tertiary and/or postgraduate education (n = 359). Asked to self-report on health, over 73.6 percent of participants rated their health as ‘good’ or ‘very good’ (n = 513), and under 26.4 percent rated it ‘poor’ or ‘very poor’ (n = 184).

The second phase of data gathering spanned the period 10^th^ October–30^th^ December 2020, when the second pandemic wave was observed in Poland. A further email was sent to respondents from the first phase, 414 of whom agreed to participate in the second round (response rate = 59.4%): 152 in the 18–35-year age group, 137 in the 36–59-year age group, and 125 in the ≥ 60 age group. Hence, respondents were grouped into three groups: younger (36.7%), middle-aged (33.1%), and older (30.2%). The mean lapse of time between the two phases was 178.04 days (SD = 25.70). There were no differences in respect of gender, self-rated health, or educational attainment between the respondents who participated only in the first phase and those who participated in both phases.

### Measures and procedure

Respondents were informed of the purpose and scope of the study, and all provided informed consent before registering for participation. Participation was voluntary and anonymous. Data were collected in an online study administered via a tool for online surveys: *www.soscisurvey.com*. Before the individuals could send their answers over the net, the data were checked for completeness. The measures outlined below were taken in both phases of data collection. Study was approved by the appropriate ethics review committee of the University of Economics and Human Sciences, prior to initiation.

### Emotional responses

Respondents were asked to rate the intensity of ‘sadness’, ‘fear’, ‘anger’, and ‘shock’ experienced due to the outbreak of COVID-19 on a 5-point scale where 1=no such emotion and 5=maximum intensity. Items were presented in random order, and the ratings given were then cross-validated using a qualitative measure. Specifically, during the first phase respondents completed the quantitative survey and were then asked an open-ended question concerning their first emotional reaction to the outbreak. The specific emotions mentioned in their responses were then coded by two independent coders (interrater reliability range 0.77-0.92). Very little difference was seen in the patterns of findings from qualitative and quantitative measures. Hence, and given that the second phase administered only the quantitative measure, only the ratings from the quantitative measure were considered in the following analysis.

### Coping strategies

To measure how respondents coped with the outbreak of the pandemic, the Brief COPE scale was used (Carver, 1997). The instrument comprised 14 two-item subscales (self-distraction, active coping, denial, substance use, use of emotional support, use of instrumental support, behavioral disengagement, venting, positive reframing, planning, humor, acceptance, religion, self-blame). Respondents were asked to rate frequency of use of the strategy outlined in each item to describe how they coped with the COVID-19 outbreak, using a 5-point scale where 1 = never and 5 = always. Hence, the higher the score, the higher the level of coping. Thereafter, we categorized the items under *problem-focused coping* (active coping, planning, use of instrumental support) or *emotion-focused coping* (use of emotional support, acceptance, positive reframing, religion, humor, substance use, self-distraction, self-blame, denial, behavior disengagement, venting; Carver, 1997). Cronbach’s alpha was used to assess the inter-item reliability of both categories, yielding a score of 0.62 for problem-focused coping and 0.74 for emotion-focused coping. Finally, the mean of all related items was used to compute a composite score for each category.

### Statistical analyses

Repeated-measures analyses of variance (ANOVAs) were performed to detect differences across age groups in relation to emotional responses shown and coping strategies deployed to counter the outbreak of the COVID-19 pandemic. Linear regression analyses were carried out to investigate the relationship between emotion and coping by assessing whether level of coping shown during the first phase could determine changes seen in emotional response between that and the second phase, or vice versa.

## Results

### Age and emotional reactions

Age-related differences in the intensity of sadness, fear, anger, and shock felt by the three cohorts in both phases were tested through an age (between-subject) x emotion (within-subject) x phase (within-subject) ANOVA. Although the three-way interaction was found to be not significant, the age-emotion interaction, in contrast, proved significant. To shed further light on these interactions, one-way ANOVAs were then carried out to investigate age difference per emotion per phase, as summarized in Table 1.

**Table 1.**
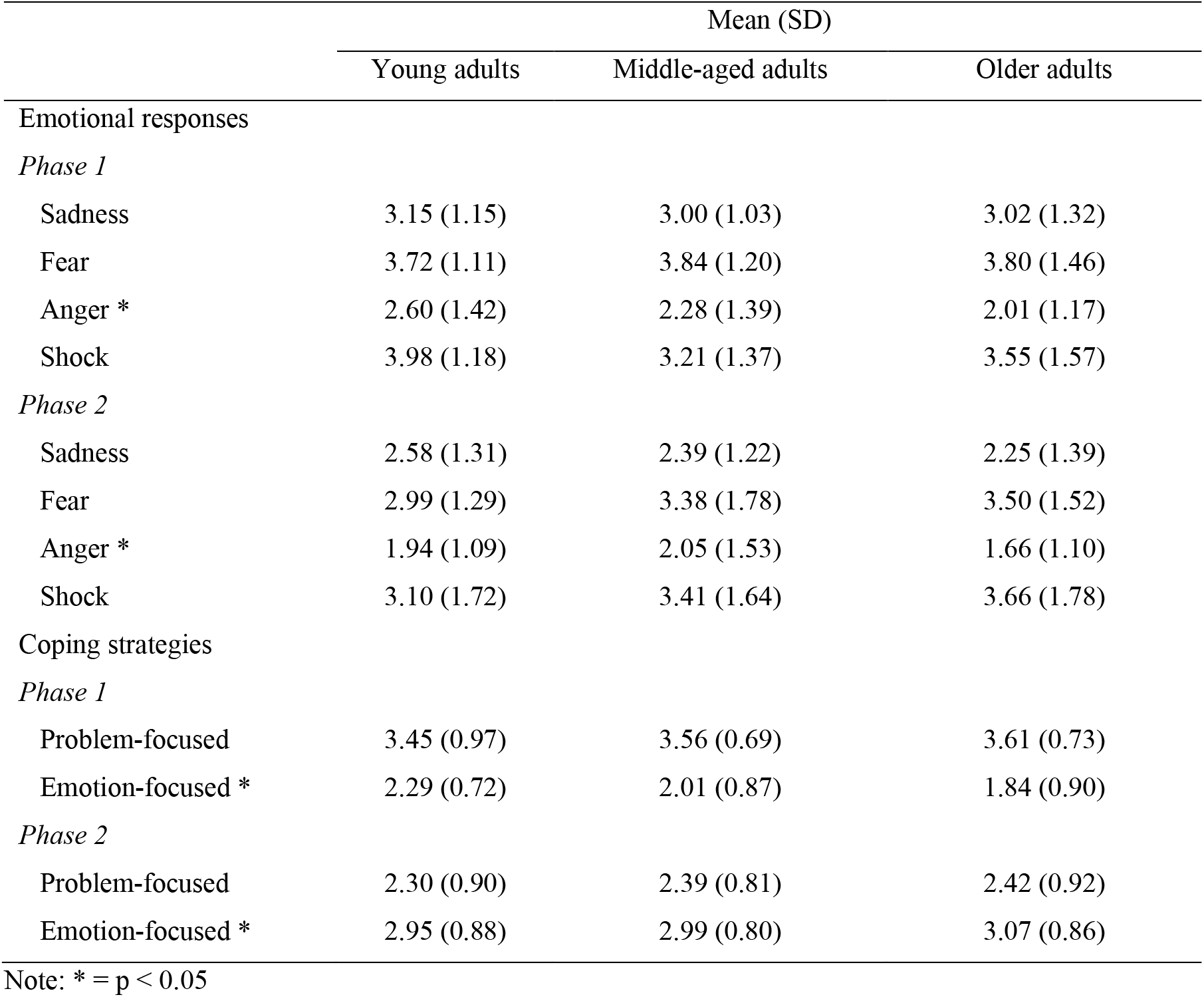
Age differences in emotions and comping strategies toward COVID-19

|  | Mean (SD) |  |  |
| --- | --- | --- | --- |
|  | Young adults | Middle-aged adults | Older adults |
| Emotional responses |  |  |  |
| *Phase 1* |  |  |  |
| Sadness | 3.15 (1.15) | 3.00 (1.03) | 3.02 (1.32) |
| Fear | 3.72 (1.11) | 3.84 (1.20) | 3.80 (1.46) |
| Anger * | 2.60 (1.42) | 2.28 (1.39) | 2.01 (1.17) |
| Shock | 3.98 (1.18) | 3.21 (1.37) | 3.55 (1.57) |
| *Phase 2* |  |  |  |
| Sadness | 2.58 (1.31) | 2.39 (1.22) | 2.25 (1.39) |
| Fear | 2.99 (1.29) | 3.38 (1.78) | 3.50 (1.52) |
| Anger * | 1.94 (1.09) | 2.05 (1.53) | 1.66 (1.10) |
| Shock | 3.10 (1.72) | 3.41 (1.64) | 3.66 (1.78) |
| Coping strategies |  |  |  |
| *Phase 1* |  |  |  |
| Problem-focused | 3.45 (0.97) | 3.56 (0.69) | 3.61 (0.73) |
| Emotion-focused * | 2.29 (0.72) | 2.01 (0.87) | 1.84 (0.90) |
| *Phase 2* |  |  |  |
| Problem-focused | 2.30 (0.90) | 2.39 (0.81) | 2.42 (0.92) |
| Emotion-focused * | 2.95 (0.88) | 2.99 (0.80) | 3.07 (0.86) |

Note: \* = $p < 0.05$

No significant age difference was detected in regard to intensity of shock, sadness, or fear in either phase; however, intensity of anger differed across the three cohorts in both phases. Post-hoc Tukey analyses were conducted, indicating that older adults experienced less intense anger in both phases, younger ones experienced the most intense levels in the first phase, and middle-aged ones experienced the most in the second phase. These findings align with our prediction that of anger is less intense among older people than younger ones when confronted with a crisis.

### Age and use of different coping strategies

Table 1 presents a summary of mean scores in both categories of coping strategies across all three cohorts and both phases. Findings of an ANOVA performed on age (between-subject) coping (within-subject) x phase (within-subject) indicated that the three-way interaction is significant. The nature of the interaction was further investigated through a one-way ANOVAs focused on age difference per strategy per phase. Findings suggested no difference across cohorts in regard to use of problem-focused coping in either phase but, in contrast, significant difference in regard to emotion-focused coping between phases. Post-hoc Tukey analyses further indicated higher frequency of emotion-based coping strategies reported by younger adults than by the other two cohorts in the first phase, but higher frequency of the same among older adults than younger ones during the second phase (when the pandemic was over 6 months present). During the second phase, no difference was found between the middle-aged cohort and the other two groups regarding emotion-focused coping.

### The relationship between emotions and coping

Carrying out a longitudinal study allowed us to investigate the relationship between coping strategies and emotional responses. Linear regression analyses were performed to determine whether changes between first and second phase in the scores for each emotion were explicable by level of coping during the first phase, or whether the reverse was true. Moreover, we investigated age-related variation in these relations. First, we tested how coping strategies influenced emotions by carrying out a regression analysis in which predictors for each change in emotion score were first, category of coping during the first phase; second, age; and third, category of coping during the second phase. Table 2 presents a summary of results. As can be seen, using problem-focused during the first phase is not a significant predictor of change in the scores for fear, anger, or shock but, in contrast, it predicts change in the score for sadness. Moreover, prediction of change in the score for sadness was also subject to a significant interaction effect between problem-focused coping and age.

**Table 2.** Regression analyses on emotion change scores toward COVID-19 as a function of coping strategies and age.

|  | Standardized beta |  |  |  |
| --- | --- | --- | --- | --- |
|  | Sadness change<br>score<sup>a</sup> | Fear change<br>score<sup>a</sup> | Anger change<br>score<sup>a</sup> | Shock change<br>score<sup>a</sup> |
| *F* (*R*<sup>2</sup>) | 18.45 (0.066)** | 3.80 (0.017) | 1.88 (0.009) | 3.42 (0.015) |
| Problem-focused coping at Phase 1 | -0.62** | -0.12 | -0.05 | -0.14 |
| Young vs old (D1) | -0.01 | -0.05 | -0.02 | 0.03 |
| Middle-age vs old (D2) | 0.02 | -0.03 | 0.01 | 0.02 |
| D1 x Problem-focused coping at Phase 1 | 0.28** | -0.01 | 0.04 | 0.01 |
| D2 x Problem-focused coping at Phase 1 | 0.22** | 0.04 | 0.05 | 0.02 |
| *F* (*R*<sup>2</sup>) | 22.35 (0.089)** | 3.12 (0.016) | 9.81 (0.077)** | 6.55 (0.055) |
| Emotion-focused coping at Phase 1 | -0.67** | -0.02 | -0.27** | -0.12 |
| Young vs old (D1) | 0.09 | -0.06 | 0.07 | 0.09 |
| Middle-age vs old (D2) | 0.07 | -0.02 | 0.09 | 0.04 |
| D1 x Emotion-focused coping at Phase 1 | 0.35** | 0.02 | -0.05 | -0.14 |
| D2 x Emotion-focused coping at Phase 1 | 0.11 | -0.01 | -0.02 | -0.12 |

Note: \* = $p < 0.05$ ; \*\* = $p < 0.001$
a: Change score = Phase 2 coping score – Phase 1 coping score.

Correlation analyses subsequently performed for each cohort revealed that older adults who had chosen more problem-focused coping strategies in the first phase experienced a greater decline in sadness over the whole period. This correlation pattern was not seen in the other two cohorts. The adoption of emotion-focused coping strategies during the first phase did not significantly predict any change in scores for fear or shock; in contrast, however, it significantly predicted a change in the score for sadness. Moreover, a significant interaction between age and emotion-focused coping qualified this effect. Across all cohorts, the tendency to use emotion-focused coping strategies was associated with a decline in the intensity of sadness between the first phase and the second. The decline was greater in older adults than among younger ones, while in regard to middle-aged adults, the correlation pattern had no more than marginal significance. Moreover, a tendency to deploy emotion-focused coping strategies during the first phase proved a significant predictor of changes in the anger score. Hence, it can be inferred that using emotion-focused coping strategies brought about a decline in anger levels across all age groups during the period.

We then carried out regression analyses (see Table 3) to determine how emotions experienced during the first phase influenced the score of each coping change across phases. Predictors were one emotion in the first phase, age, and age x emotion in the first phase. None of the four emotions experienced during the first phase predicted the change in scores for either problem-focused or emotion-focused coping strategies. In contrast, significant interaction was noted between fear and age in regard to the change in score for problem-focused coping. A correlation analysis was then conducted for each cohort, which indicated that those members of the youngest cohort who were more worried about COVID-19 during the first phase saw a greater decline in problem-focused coping over the whole period, while this correlation was not significant for the two older cohorts. To sum up, the findings of the regression analyses indicated that, during the first phase, coping was a better predictor of emotions than vice versa. Across all three cohorts, a tendency to use emotion-focused strategies during the first phase was associated with lower levels of anger and sadness, while a tendency to use problem-focused strategies led to less sadness among older adults. On the other hand, emotions experienced during the first phase did not, on the whole, serve as predictors for changes in coping strategy overt the whole period, except that a greater decrease in problem-focused coping was seen among younger adults who exhibited more intense feelings of worry about COVID-19 during the first phase.

**Table 3.**
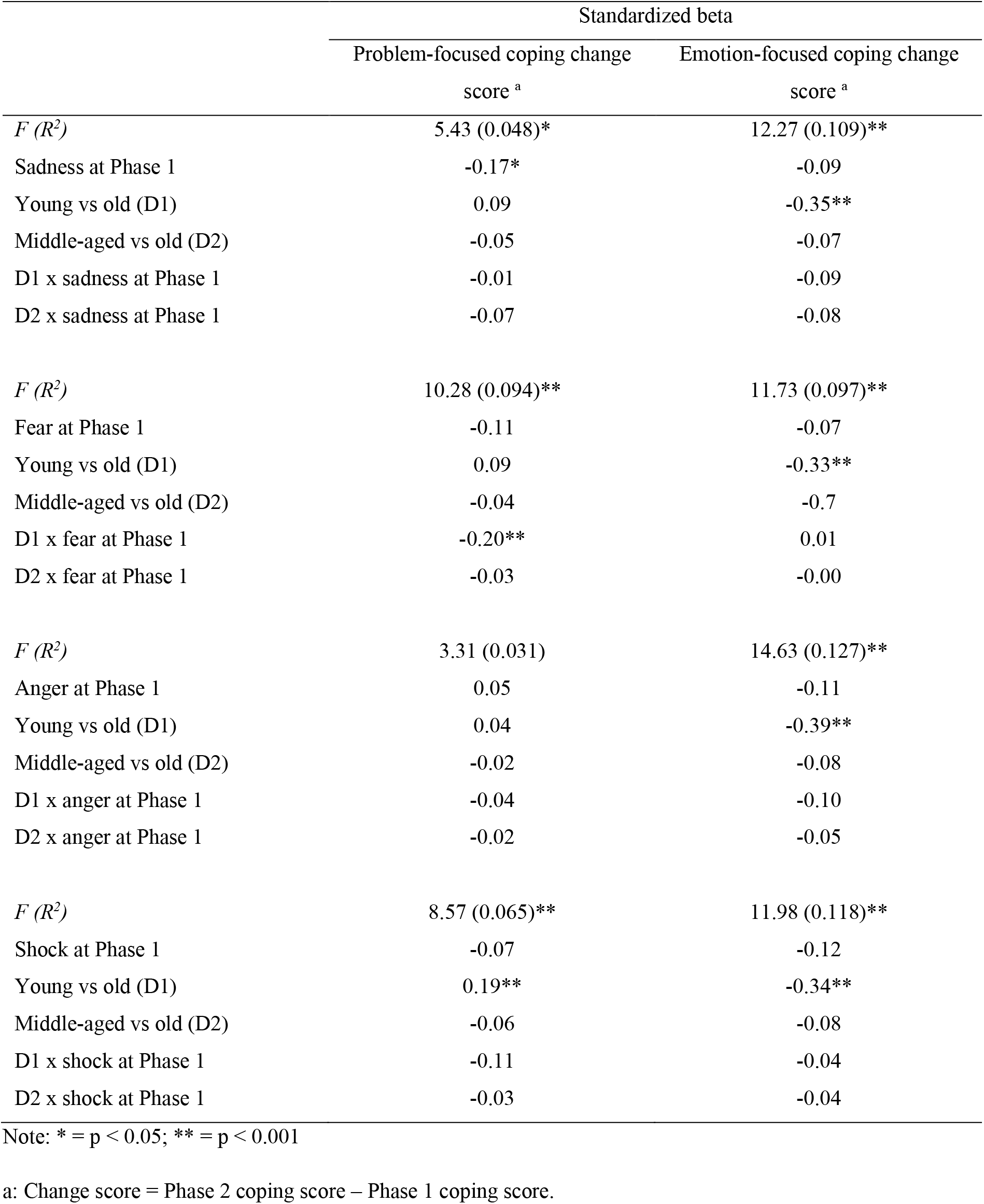
Regression analyses on coping strategy change scores toward COVID-19 as a function of emotions and age.

## Discussion

The current study used the opportunity presented by the COVID-19 pandemic to investigate whether emotional responses and coping strategies vary across age groups. We anticipated that our own findings would replicate those of prior research on age-related emotions and coping in response to a crisis, namely that older people experience more sadness than young ones, but less anger and fear and, moreover, demonstrate greater use of emotion-focused coping strategies and less of problem-focused ones. Furthermore, we have bridged a gap in the literature through a longitudinal study of the relationship between age-related emotions and coping strategies.

### Age and emotional reactions

The findings partially confirm our prediction of relations between age difference and emotion. While all three cohorts felt more intense sadness, anger, shock, and fear during the first phase than the second (i.e., at the first peak rather than when the virus was somehow known), the lowest levels of anger were shown by older adults in both phases. Hence, it can be inferred that, in a crisis situation, older adults are consistently less likely to experience anger than younger ones. These findings align with prior research into age-related difference in emotion in regard to other types of crisis (Charles et al., 2001; Felton & Revenson, 1987; Knight et al., 2000; Weiner & Graham, 1989). Moreover, they support the proposition of Charles and colleagues (2001) that, when confronted by a problematic situation, younger adults will more likely have an active, solution-oriented emotional response than older ones. As regards shock, sadness, and fear, however, our results indicate no significant age-related difference over time. It must be acknowledged that these null effects may be due to the particular characteristics of the COVID-19 pandemic, which has been unprecedented in terms of numbers affected, whether directly or indirectly, as well as lack of pre-existing therapy. In this situation, shock, fear, and sadness can be considered normative emotional responses across all age groups.

### Age and use of different coping strategies

All three cohorts made less frequent use of problem-focused coping strategies (e.g., active coping, use of instrumental support) and more frequent use of emotion-focused ones (e.g., use of emotional support, humor, behavior disengagement) from one phase to another. This may indicate they felt less need to find solutions and instead privileged the need to move on emotionally as the pandemic was over 6 months present. Within this overall trend, interesting variations were seen. During the first phase, younger adults were more likely to use emotion-focused coping strategies than middle-aged and older ones. All three cohorts made more frequent use of emotion-focused coping strategies as the disease was controlled, but middle-aged and older adults did so to a greater extent than younger ones. As a result, during the second phase, the age difference in regard to type of coping strategy adopted had been reversed.

If we regard changes in strategy adopted between the two phases as a coping process and consider only the end point of this process, it can be seen that the oldest age group used more emotion-focused coping strategies than the younger ones. This finding bears out our prediction regarding age differences in coping strategies and replicates the results of previous laboratory studies of age-related emotion-focused coping, suggesting a good level of external validity (Blanchard-Fields et al., 1995; Charles et al., 2001). However, if we consider the pattern of our findings overall, it appears that time of measurement is key in the investigation of such differences as assessing the strategies adopted at different periods of the overall coping process can show different patterns in how age groups react. It has been suggested that the occasional inconsistency in the results of previous research in this area is due to older adults being in possession of a wider range of coping strategies, which, moreover, they are better equipped to deploy flexibly according to context (Aldwin, Sutton, Chiara, & Spiro, 1996; Prince-Embury & Rooney, 1990). Such inconsistency may also be due to which type of emotion-focused strategy, exactly, is being measured. However, the results of the current study indicate that this inconsistency may also arise from the fact that coping strategies change over time; thus, measuring them at one moment only may not reveal a full picture of the coping process (Blanchard-Fields et al., 1995, 1997; Blanchard-Fields, Stein, & Watson, 2004).

This study found no difference across age groups in regard to problem-focused coping strategies, possibly due to the specific characteristics of COVID-19, as outlined above. There are few problem-focused (active coping) strategies available to individuals confronting COVID-19, and they constitute a well-known list which allows little room for age-related difference: observing hand hygiene, wearing facemasks, using instrumental support, and seeking information and advice (Shanahan et al., 2020; Trnka & Lorencova, 2020). Again, these findings chime with previous studies carried out in instrumental settings, which also found few age-related differences in regard to problem-solving coping strategies (Blanchard-Fields et al., 1995, 2004).

### The relationship between emotions and coping

The most important contribution made by our longitudinal analyses is to determine the flow of the cycle linking age-related emotions and coping: it appears that emotions arise from coping strategies more than vice versa. Hence, individuals of all ages who made more use of emotion-focused coping in the first phase were likely to experience less anger and sadness throughout the pandemic, while older adults who made more use of problem-focused coping experienced less sadness. In contrast, changes to coping strategy during the pandemic could not, in general, be predicted by the emotional responses reported when the outbreak was at its worst, with the exception that problem-focused coping decreased more among younger adults who initially experienced higher levels of worry. These results are in alignment with the coping literature and indicate that emotions are more dependent on coping strategies, in particular emotion-focused ones, than vice versa (Folkman et al., 1987). Although certain emotions may have specific motivational properties, our results indicate they do not give rise to specific coping strategies, at least in response to the COVID-19 pandemic.

## Conclusions and limitations

Overall, the age-related emotional differences and coping strategies recorded during this research suggest that older adults dealt with the outbreak of COVID-19 better than younger ones, experiencing both less anger during the period covered and a higher decrease of negative emotions due to their use of both types of coping strategy. Our findings are in accordance with a general consensus in the literature that the older people become, the better equipped they are to regulate their emotions (Carstensen, Fung, & Charles, 2003). This increased ability to regulate emotion may explain why robust emotional well-being can be maintained even during the difficulties of the aging process (Charles, Reynolds, & Gatz, 2001).

Certain limitations on the present study must be acknowledged. First, this study directly measured neither perceived controllability nor dispositional factors which could influence emotional state and choice of coping strategy in the face of the pandemic. These limitations are mitigated by both the longitudinal nature of our research and the fact that COVID-19 had an impact on people of all ages and types; however, the possibility remains that such variables may confound some of our findings. Second, the cross-sectional nature of this investigation precludes us from drawing causal inferences. Third, although Internet-based data collection can provide substantial statistical power, the sample of participants may be biased toward those individuals who have access to and regularly use the Internet. Fourth, we relied exclusively on self-report measures.

## Data Availability

The raw data of this study are available from the corresponding author upon reasonable request.

## Data

The raw data of this study are available from the corresponding author upon reasonable request.

